# Impact of sampling site on diagnostic test accuracy of RT-PCR in diagnosing Severe Acute Respiratory Syndrome Coronavirus 2 (SARS-CoV-2) infection since the emergence of omicron: a systematic review and meta-analysis

**DOI:** 10.1101/2023.10.09.23296728

**Authors:** Lena Saal-Bauernschubert, Carina Wagner, Alexey Fomenko, Theo Dähne, Ana-Mihaela Bora, Heidrun Janka, Stephanie Stangl, Nicole Skoetz, Stephanie Weibel

## Abstract

Nasopharyngeal sampling (NP) is the routine standard for SASR-CoV-2 detection using reverse transcription polymerase chain reaction (RT-PCR). In this systematic review, we assessed diagnostic test accuracy of alternative sampling sites compared to NP for RT-PCR testing of Omicron (sub)-variants.

We systematically searched for studies from January 2022 until February 2023 investigating any type of respiratory sample for RT-PCR in people with suspected, known, or known absence of SARS-CoV-2 Omicron infection. Data were pooled for each comparison using the bivariate model, sensitivity and specificity was estimated with 95% confidence intervals (CIs). Risk of bias was assessed with QUADAS-2 tool, certainty of evidence with GRADE.

We included three cohort-type cross-sectional studies (1,003 participants). Saliva versus NP sampling in three studies showed a sensitivity of 92% (95% CI 87% to 96%) and a specificity of 94% (95% CI 83% to 98%). AN versus NP sampling in one study showed a sensitivity of 90% (95% CI 82% to 95%) and a specificity of 99% (95% CI 95% to 100%). Certainty of evidence for sensitivity and specificity of both comparisons was low to very low.

Based on the current very low- to low-certainty evidence, we are uncertain about accuracy of different sampling sites for RT-PCR.

## 1 INTRODUCTION

The WHO announced on May 5^th^, 2023 that COVID-19 would no longer be considered a public health emergency of international concern (PHEIC) heralded a new phase of the ongoing pandemic. By that time, the virus had affected over 765 million people and resulted in the loss of over 6.9 million lives.^1,2^ Notwithstanding the WHO announcement, the danger of emerging variants and subsequent outbreaks persists, underscoring the continued importance of maintaining the testing strategies.^3^ Over the course of the pandemic, marked by various surges of viral variants, reverse transcription polymerase chain reaction (RT-PCR) remained an important cornerstone for the diagnosis of SARS-CoV-2 infections.

The prevailing virus variant, Omicron, exhibits an extensive number of over 60 mutations, resulting in modified transmission dynamics that contribute to its rapid spread and pose the potential of altered diagnostic test accuracy (DTA).^4^

Additionally, diagnostic test accuracy varies depending on type of upper respiratory specimen used. Diagnostic tests for the detection of SARS-CoV-2 via RT-PCR are often only validated for one particular type of upper respiratory specimen. This results in uncertainty of accuracy and consequently problems in interpretation of test results when deviating from the validated type of specimen. However, deviations can be necessary because of a multitude of factors including (but not limited to) anatomical changes due to prior surgery, bleeding disorders or insufficient compliance because collection of a particular specimen is experienced as unpleasant. For outbreak scenarios rapid actions are crucial in order to limit the spread of a disease. Here it can be advantageous to deviate from the validated specimen and use a more easily and quickly obtainable specimen like saliva instead of a nasopharyngeal swab. This should only be done with comprehensive knowledge of possible concomitant limitations regarding the impact of sampling site on test accuracy. To our knowledge, there is a lack of systematic investigation of the accuracy of RT-PCR tests in diagnosing SARS-CoV-2 infection during the Omicron period, particularly in relation to the specific respiratory specimen employed. Therefore, we set out to gather available evidence on the impact of the sampling site on test accuracy.

The present meta-analysis is part of the upgrade of the German AWMF-S1 guideline to a German AWMF-S3 guideline for the importance of RT-PCR testing of health care workers.^5^

## 2 MATERIALS AND METHODS

We developed a study protocol following standard guidelines for systematic reviews.^6,7^ The protocol for this systematic review was registered on the International prospective register of systematic reviews (PROSPERO, identifier CRD42023416220) and made publicly available on April 11^th^ 2023.

### Eligibility criteria

#### Types of studies

We considered primary studies, which allowed comparisons between different sampling sites used to diagnose SARS-CoV-2 infection. We included studies of all designs that produced estimates of test accuracy or provided data from which we could compute estimates. We considered cohort type cross-sectional studies, which recruited participants before disease status had been ascertained and case-control cross-sectional studies, where individuals were recruited based on the knowledge of the presence or absence of the target.^8^ Only studies reported as full text journal publications were eligible. We excluded studies from which we could not extract data to compute sensitivity and specificity. Studies with inconsistencies regarding data were hold in awaiting classification and study authors were contacted. Preprints, conference abstracts, letters, editorials and ongoing studies were excluded. Only studies in English language were included.

#### Types of participants

We included studies recruiting people presenting with suspicion of current SARS-CoV- 2 infection (e.g., symptomatic or close contacts) or those recruiting people who were screened for infection independent of symptoms and history (i.e., hospital or community screening). We included all studies irrespective of the investigated population (i.e., health care workers (HCWs), in- and outpatients, general population). We restricted the review to studies recruiting people or parts of the study participants (e.g., presented as subgroup analysis) during an omicron wave or later (i.e., December 2021 onwards).

#### Types of index test and reference test

We included any available RT-PCR for diagnosis of SARS-CoV-2. Any respiratory sampling site and strategies based on multiple applications (i.e., combined nasopharyngeal and oropharyngeal sampling) of a test were also eligible for inclusion. We considered nasopharyngeal (NP) sampling as the reference sampling site in the assessment of test accuracy of RT-PCR sampling sites.

#### Types of outcome measures

We analysed the following outcomes: diagnostic test accuracy, including sensitivity and specificity, positive predictive value (PPV) and negative predictive value (NPV). For quality assurance True Positives (TP), True Negatives (TN), False Positives (FP), and False Negatives (FN) have been extracted (if provided) and all outcomes regarding test accuracy have been recalculated (i.e., calculated sensitivity, calculated specificity etc.) to confirm findings. Furthermore, Positive and Negative Likelihood Ratios as well as Diagnostic Odds Ratio have been calculated. Likelihood ratios are independent of disease prevalence. The interpretation of likelihood ratios is intuitive: the larger the positive likelihood ratio, the greater the likelihood of disease; and vice versa^9^.

### Search strategy

The literature was searched by an experienced information specialist (HJ) from 1st January 2022 until 8th February 2023 using four bibliographic databases: PubMed, Cochrane COVID-19 Study Register (covid19.cochrane.org), Science Citation Index Expanded (Clarivate Web of Science) and the WHO COVID-19 Research database (search.bvsalud.org/global-literature-on-novel-coronavirus-2019-ncov/). References of included studies were screened to identify additional records. The time limit was applied to retrieve literature published on the variant Omicron. No language restrictions were applied. Details of the search strategies are available as supplementary material (Supplemental Material Table S1).

### Selection of studies and data extraction

#### Selection of studies

Duplicate records were removed using an automated artificial intelligence (AI)-based deduplication solution, Deduklick.^10^

Two review authors independently screened the retrieved titles and abstracts for potentially eligible studies using Covidence software (https://www.covidence.org/). Any disagreement regarding title and abstract screening was dissolved by discussion or by consulting a third review author if necessary.

Two review authors then independently assessed the full-text articles of the selected studies against the inclusion criteria. Any disagreement was resolved by discussion or by consulting a third reviewer.

When more than one article presented data on the same population, we included the primary article, which would be the article with the largest number of people or with the most informative data.

When a study did not present all relevant data, we contacted the study authors directly to request further information.

#### Data extraction

Two review authors independently performed data extraction. Any disagreements regarding data extraction were dissolved by discussion or by consulting a third review author if necessary. We extracted data on study characteristics (study design, study location, recruitment period), details on study participants, index test, and reference test, flow and timing, and outcomes.

### Assessment of methodological quality in included studies

The risk of bias and methodological quality of each included study was assessed using the Quality Assessment of Diagnostic Accuracy Studies 2 tool (QUADAS-2),^11^ as recommended by Cochrane. This tool consists of four domains: patient selection, index test, reference standard, flow and timing. For each domain, the risk of bias was analysed using different signalling questions, which were answered either by yes/no or by low/high/unclear risk of bias or by low/high/unclear concern. The tool was tailored according to this review question (Supplemental Material Table S2).

Two review authors independently performed the QUADAS-2 assessments. Any disagreement was resolved by discussion or by consulting a third review author. Each domain was assessed in terms of risk of bias, and the first three domains were also considered in terms of applicability concerns.

### Data synthesis

The meta-analyses were performed using the MetaDTA tool (https://crsu.shinyapps.io/dta_ma/). ^12,13^

We included studies based on either participants or samples. We collected counts on TP, TN, FP and FN test results per study (samples or participants) to construct a two-by-two table and estimated sensitivity and specificity with 95% confidence intervals. We plotted the sensitivities and specificities with their respective 95% CI in forest plots. Overall sensitivity and specificity were calculated using a random effects binominal bivariate analysis fitted as a generalised linear mixed effect model.^12^ PPV and NPV have been calculated using the information on Positive and Negative Likelihood Ratios derived from the MetaDTA output according to the following formulas:

PPV=FEST(((prevalence/(1-prevalence))*LR+)/((prevalence/(1-prevalence))*LR+ + 1);2)

NPV=FEST(1-((prevalence/(1-prevalence))*LR−)/((prevalence/(1-prevalence))*LR− + 1);2)

PPV and NPV have been calculated for a prevalence of 0.0279% which was the prevalence of SARS-CoV-2 infections in May 2023.^14^

We planned not to perform meta-analysis in case of too large clinical or methodological heterogeneity. However, studies were considered homogeneous allowing pooling. We investigated heterogeneity between studies by visually inspecting the forest plots of sensitivity and specificity. Non-overlapping 95% CIs were considered as heterogeneous study results.

### Analysis of subgroups or subsets

We predefined subgroups for meta-analysis based on the following characteristics: sample collection (professional or layman), sampling site of the index test, testing procedure in accordance to manufacturer’s instructions, presence or absence of symptoms, sampling time after symptom onset (early = up to seven days, late ≥ seven days), population, Cycle threshold (Ct) value cut-off.

Subgroup analyses were not performed due to insufficient number of studies.

### Sensitivity analysis

A sensitivity analysis was conducted to assess the influence of risk of bias on study results.

### Summary of findings and assessment of the certainty of the evidence

The certainty of the evidence was assessed using the GRADE (Grading of Recommendations, Assessment, Development and Evaluation) approach for test accuracy.^15,16^ We used the softwareGRADEpro (https://www.gradepro.org/) to create summary of findings tables for the following outcomes: Diagnostic test accuracy (including sensitivity, specificity)

### Assessment of the certainty of the evidence

The GRADE approach uses five domains (risk of bias, inconsistency, imprecision, indirectness, and publication bias) to assess certainty in the body of evidence for each outcome.

For interpretation of absolute numbers of FP and FN, we used a pre-test probability of 0.0279% which was the prevalence of SARS-CoV-2 infections in May 2023.^14^ In light of lacking recommendations regarding the performance of diagnostic test accuracy for RT-PCR in the detection of SARS-CoV-2 we decided to establish our own threshold values to judge imprecision, adapted to WHO recommendations for Ag-RDT: based on its excellent sensitivity in the detection of SARS-CoV-2-RNA in respiratory specimens we decided to raise the lower level of acceptable sensitivity from 80% that has been recommended by WHO for Ag-RDTs to 90% for RT-PCR from any specimen used in our work. For specificity we decided to apply a threshold value of >97% as suggested for Ag-RDT by WHO.^17^

We downgraded our certainty of the evidence for:

- serious (−1) or very serious (−2) risk of bias;
- serious (−1) or very serious (−2) inconsistency;
- serious (−1) or very serious (−2) uncertainty about directness;
- serious (−1) or very serious (−2) imprecise or sparse data;
- serious (−1) or very serious (−2) probability of publication bias.

The GRADE system uses the following criteria for assigning grade of evidence.

- ‘High’: we are very confident that the true effect lies close to that of the estimate of the effect.
- ‘Moderate’: we are moderately confident in the effect estimate; the true effect is likely to be close to the estimate of effect, but there is a possibility that it is substantially different.
- ‘Low’: our confidence in the effect estimate is limited; the true effect may be substantially different from the estimate of the effect.
- ‘Very low’: we have very little confidence in the effect estimate; the true effect is likely to be substantially different from the estimate of effect.

## 3 RESULTS

### Search

We searched all databases and screened the resulting records up to 8th February 2023. Our search strategy identified a total of 11,043 records. After removal of 21 duplicate records additionally identified by Covidence, title and abstract of 11,022 records were screened by two authors. 11,016 records were excluded and six studies were assessed for eligibility. After full text screening of these six studies, three studies were included in this study. Two studies were excluded with reasons because they did not report data needed to calculate specificity.^18,19^ One study is still awaiting classification due to inconsistencies in reported data.^20^ In total, three studies are included in the qualitative and quantitative synthesis (meta-analysis).^21–23^ The PRISMA flow diagram with reasons for exclusion of studies is provided in Figure 1.

**Figure 1:**
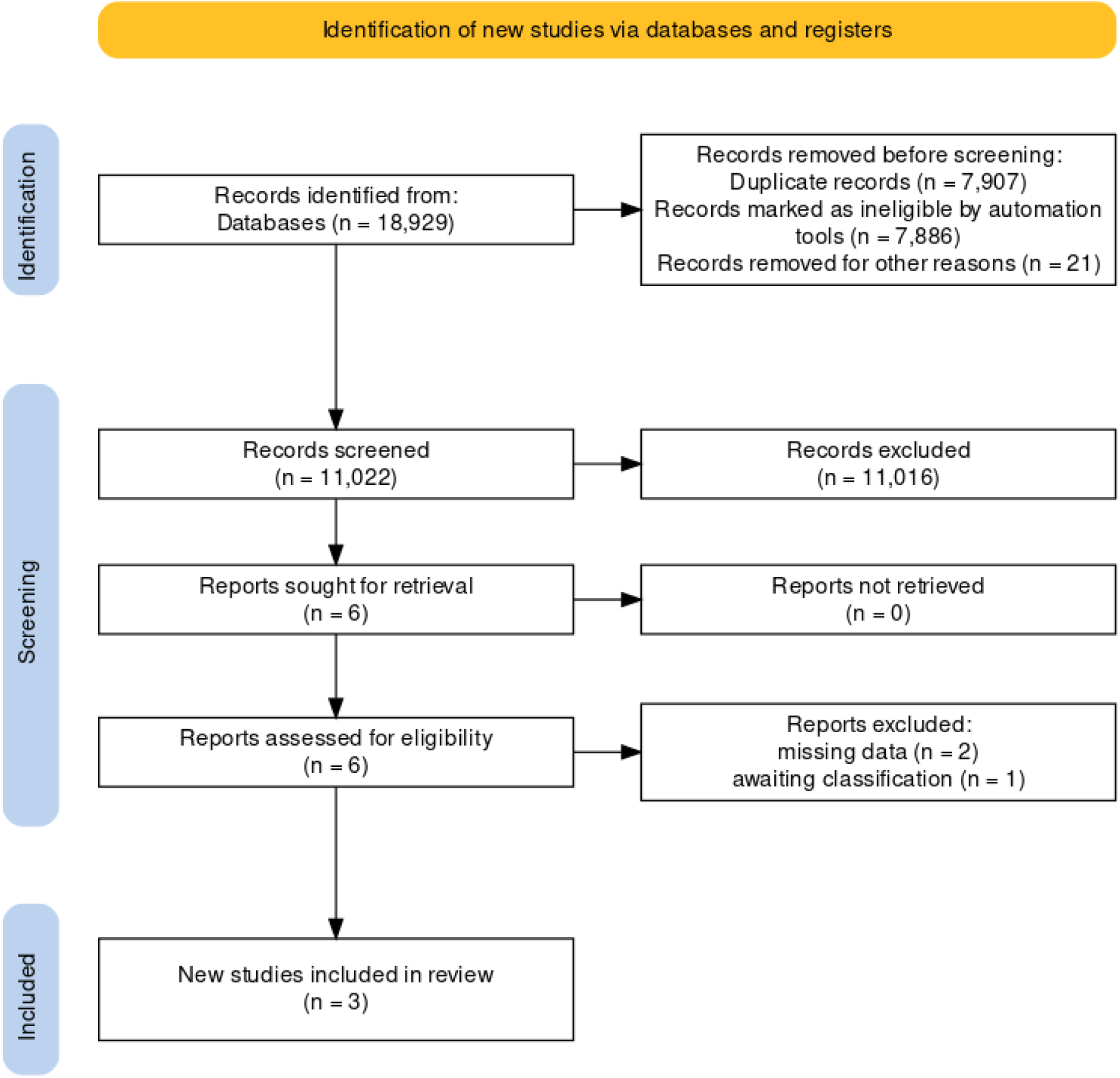
PRISMA flow diagram. Based on Haddaway et al. (2022)^31^

### Study characteristics

In total, 1,003 patients in three cohort type cross-sectional diagnostic test accuracy (DTA) studies were eligible (for individual study details, see Table 1).^21–23^

**Table 1:** Study characteristics.

| INCLUDED STUDIES |  |  |  |  |  |  |  |
| --- | --- | --- | --- | --- | --- | --- | --- |
| Study Reference | Study Design | Recruitment Period | Setting and Patient Status | N (Participants) | Index Test | Reference Test | Outcomes |
| Kritikos 2022 | cohort type cross-sectional study | 04 February - 31 March 2022 | symptomatic health care workers | 475 | RT-PCR (saliva); sampling method: swab by Health care worker/professionals; Kit: NR | RT-PCR (nasopharyngeal); sampling method: Health care worker/professionals; test platform: Cobas 6800, Roche | 2x2 table, sensitivity (positive agreement), specificity (negative agreement) |
| Migueres 2022 (saliva) | cohort type cross-sectional study | 02 February 2022 - 19 February 2022 | General population (People at risk of COVID-19 (symptomatic individuals or asymptomatic contact cases) or previously diagnosed) | 202 | RT-PCR (saliva); sampling method: saliva spitting, supervised self-collection; Kit: Thermo Fisher TaqPath COVID-19 RT-PCR kit | RT-PCR (nasopharyngeal); sampling method: Health care worker/professionals; test platform: QuantStudio 5 RT-PCR System | incomplete 2x2 table + text, sensitivity (calculated), specificity (calculated) |
| Migueres 2022 (AN) | cohort type cross-sectional study | 02 February 2022 - 19 February 2022 | General population (People at risk of COVID-19 (symptomatic individuals or asymptomatic contact cases) or previously diagnosed) | 202 | RT-PCR (AN); sampling method: swab by Health care worker/professionals; Kit: Thermo Fisher TaqPath COVID-19 RT-PCR kit | RT-PCR (nasopharyngeal); sampling method: Health care worker/professionals; test platform: QuantStudio 5 RT-PCR System | incomplete 2x2 table + text, sensitivity (calculated), specificity (calculated) |
| Ursic 2022 | cohort type<br>cross-sectional study | January 2022 | General population (People<br>visiting routine testing (not<br>further specified)) | 326 | RT-PCR (saliva);<br>sampling method:<br>saliva collector,<br>supervised self-collection; Kit: CE IVD<br>LightMix Kit SARS-CoV-2 E+N UBC | RT-PCR (nasopharyngeal);<br>sampling method: Health care<br>worker/professionals; test<br>platform: NR | 2x2 table,<br>positive<br>agreement (PPV),<br>negative<br>agreement (NPV) |

Abbreviations: RT-PCR: real-time polymerase chain reaction; NR: not reported; PPV: positive predictive value; NPV: negative predictive value

The study by Kritikos 2022^21^ was the largest study with 475 participants conducted in Switzerland. The study investigated symptomatic health care professionals (453 participants) and hospitalized patients (22 participants) with COVID-19 compatible symptoms. Two studies^22,23^ focused on patients from general population. Migueres 2022^22^ took place in France and included 202 people at risk of COVID-19 (symptomatic individuals and asymptomatic contact persons). Ursic 2022^23^ was carried out in Slovenia and included 326 people visiting a routine testing centre. All three studies^21–23^ included vaccinated people with the majority receiving at least two doses of vaccination: Kritikos 2022 84%, Migueres 2022 92%, Ursic 2022 47%.

For our analyses we created the following two comparisons each with NP sampling as reference standard: saliva versus NP sampling and AN versus NP sampling. All three studies investigated saliva versus NP sampling although the type of sampling differed. In Kritikos 2022 sampling of saliva swab and NP swab was performed by a health care worker. Migueres 2022 (saliva) used saliva spitting carried out by supervised self-collection and NP swab performed by a health care worker. In the study of Ursic 2022 saliva sampling was also performed by supervised self-collection and nasopharyngeal swab by a professional health care worker. The study by Migueres 2022 (AN) additionally analysed AN versus NP sampling. Both samples were collected with swabs by a professional health care worker.

The sampling sequence is insufficiently reported in most of the studies (Kritikos 2022, Migueres 2022 (AN), Ursic 2022). Only the study of Migueres 2022 (saliva) reports an appropriate sampling sequence according to saliva sampling first and NP sampling second.

All three studies showed their results of the saliva and NP sampling in a 2×2 table (see Table 2). Kritikos 2022 only reported sensitivity and specificity for health care workers and not for inpatients, therefore, the inpatients were excluded from our analysis. Furthermore, Kritikos 2022 reported sensitivity as positive agreement and specificity as negative agreement. Migueres 2022 did not report any specificity. The study by Ursic 2022 did not specifically report on sensitivity and specificity but also on positive agreement and negative agreement. All three studies show slight differences in reported sensitivity/specificity and calculated sensitivity/specificity.

**Table 2:**
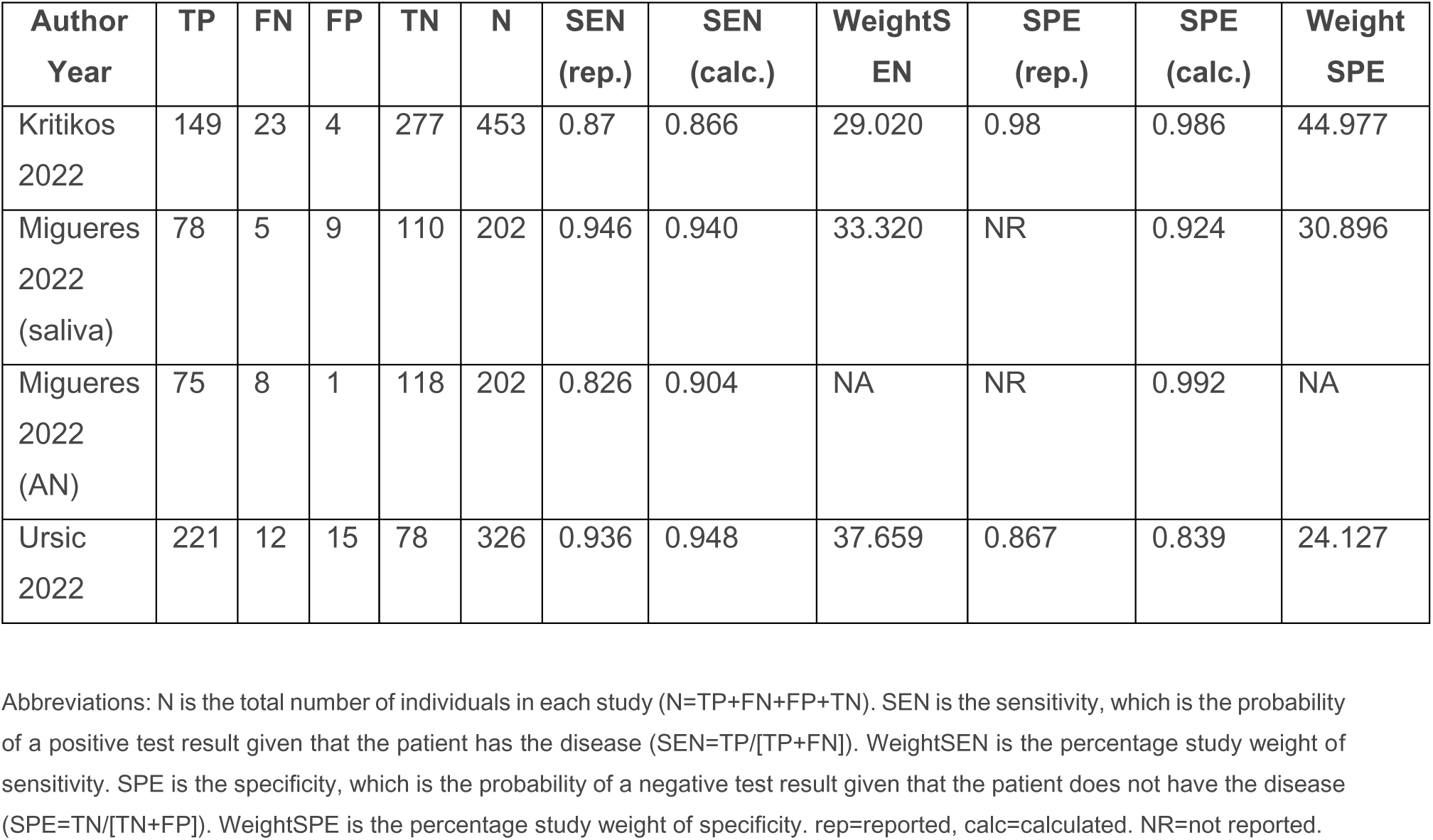
Study-level accuracy outcomes.

One identified study is not yet included in our analysis but is awaiting classification^20^. The study is a cohort type cross-sectional study, taking place in Iraq. The study included 100 symptomatic patients and analysed saliva swab versus NP swab obtained by a health care worker. Due to lack of clarification of inconsistencies in reported and calculated sensitivity and specificity, the study is not yet included in the analysis. An author request was sent.

### Risk of bias of included studies

The assessment of study quality using the QUADAS-2 tool is presented in Table 3. In all included studies, patient selection was assessed as low risk of bias as a case-control design and inappropriate exclusions were avoided. The domains index test and reference standard were assessed as unclear risk of bias in all three studies. As none of the studies reported a cut-off Ct-value for defining a positive or negative test, we assessed these domains as unclear risk of bias because of insufficient reporting. Even a blinding of the persons interpreting the particular test is not reported. However, storage conditions were rated as adequate for all studies. The domain flow and timing was also assessed with unclear risk of bias for the studies Kritikos 2022, Ursic 2022 and Migueres 2022 (AN) because the sequence of sampling was not sufficiently reported for these three studies. Only the study Migueres 2022 (saliva) reported an appropriate sequence of sampling in the domain flow and timing and was assessed as low risk of bias.

**Table 3:**
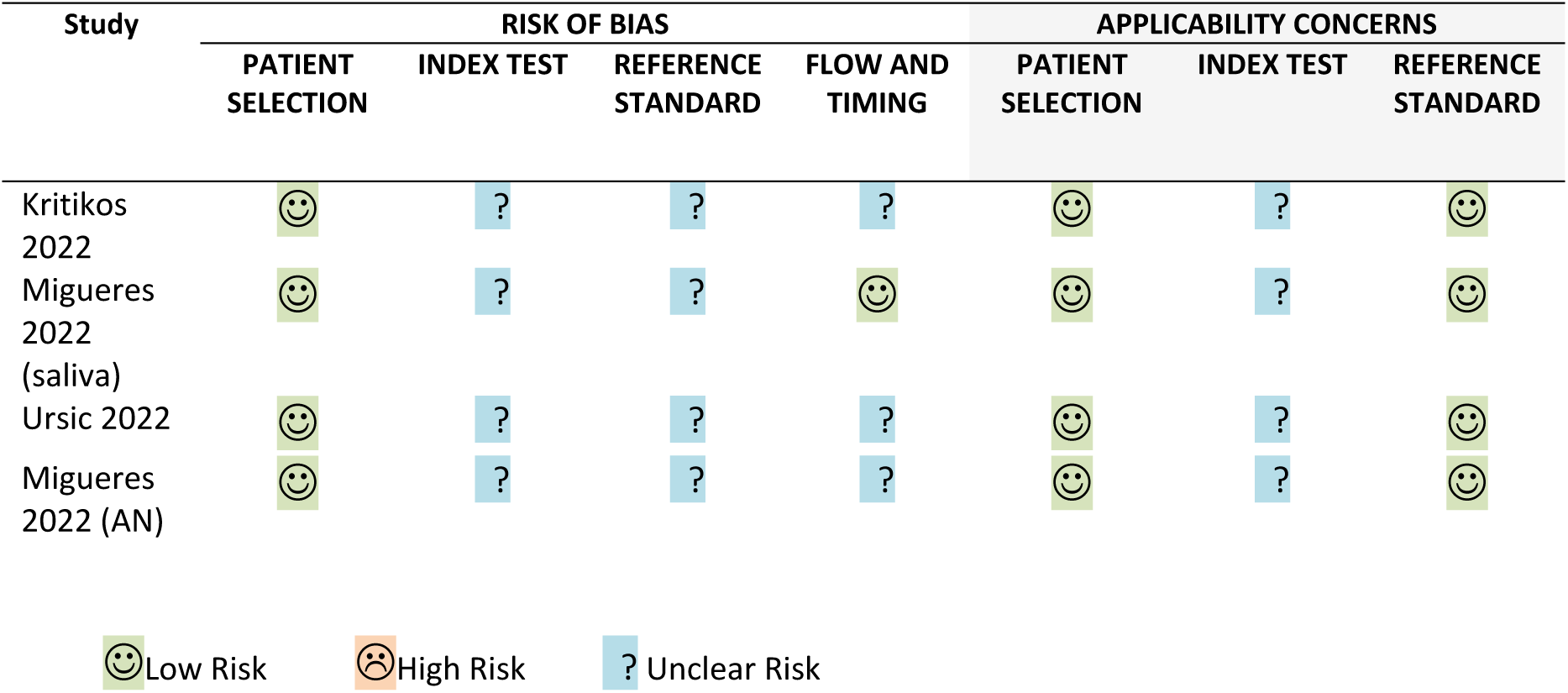
Risk of bias of included studies according to QUADAS-2.

In terms of applicability concerns, all included studies were assessed with low concern for the domains patient selection and reference standard. The domain index test was rated with unclear concern in all studies because it is not clear if the index test, its conduct or its interpretation differ from the review question and because of the insufficient reporting of a cut-off Ct-value.

### Detection of SARS-CoV-2 infection in saliva and AN sampling compared to NP sampling

Three studies were included in our meta-analysis of saliva versus NP sampling (Table 4). Across the three datasets (981 patients), the pooled sensitivity and specificity were 92% (95% CI 87% to 96%) and 94% (95% CI 83% to 98%). False positive rate was 0.058 (95% CI 0.02 to 0.17) and diagnostic odds ratio was 193.448 (95% CI 81.75 to 457.98). The positive likelihood ratio was 15.953 (95% CI 5.20 to 48.98) and the negative likelihood ratio was 0.082 (95% CI 0.05 to 0.14). For a prevalence of 0.0279% we calculated a PPV of 31% (95% CI 13%to 58%) and a NPV of 100% (95% CI 100% to 100%).

**Table 4:**
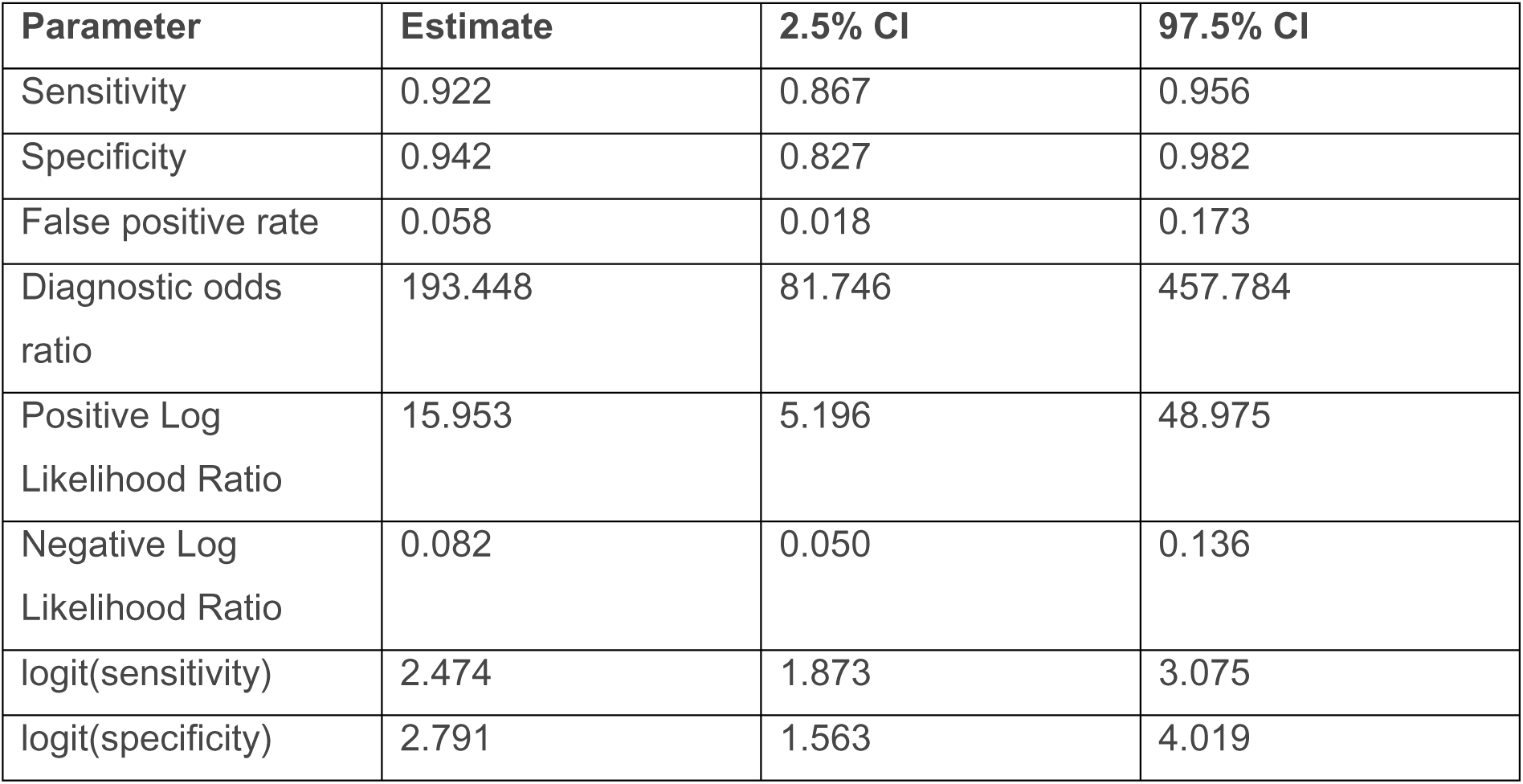
Statistic parameters of bivariate meta-analysis for the comparison of saliva versus nasopharyngeal sampling (3 studies)

Sensitivity and specificity of each study is presented in Table 2 and the forest plots in Figure 2 and 3.

**Figure 2:**
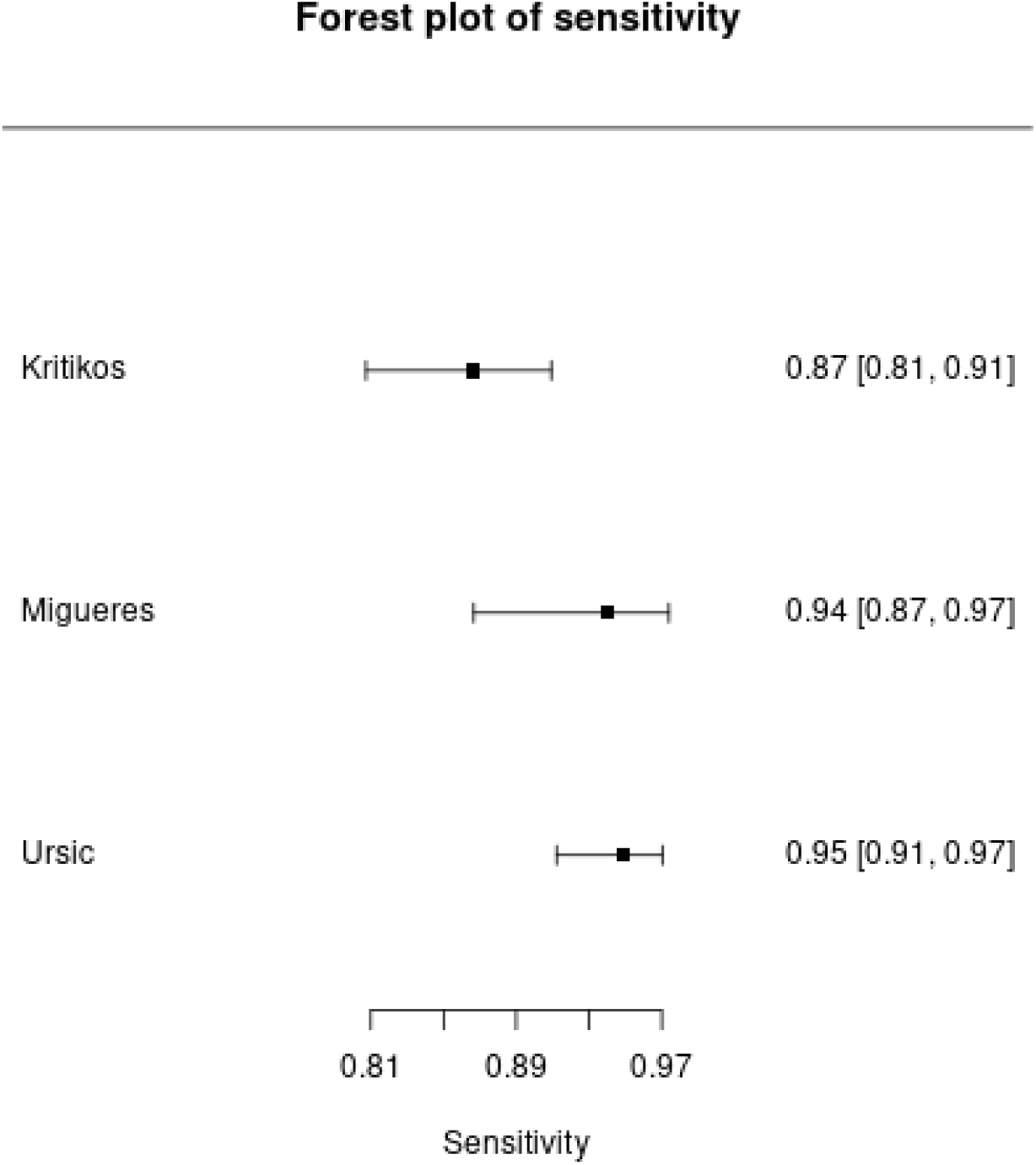
Forest plot of sensitivity for saliva versus nasopharyngeal sampling.

**Figure 3:**
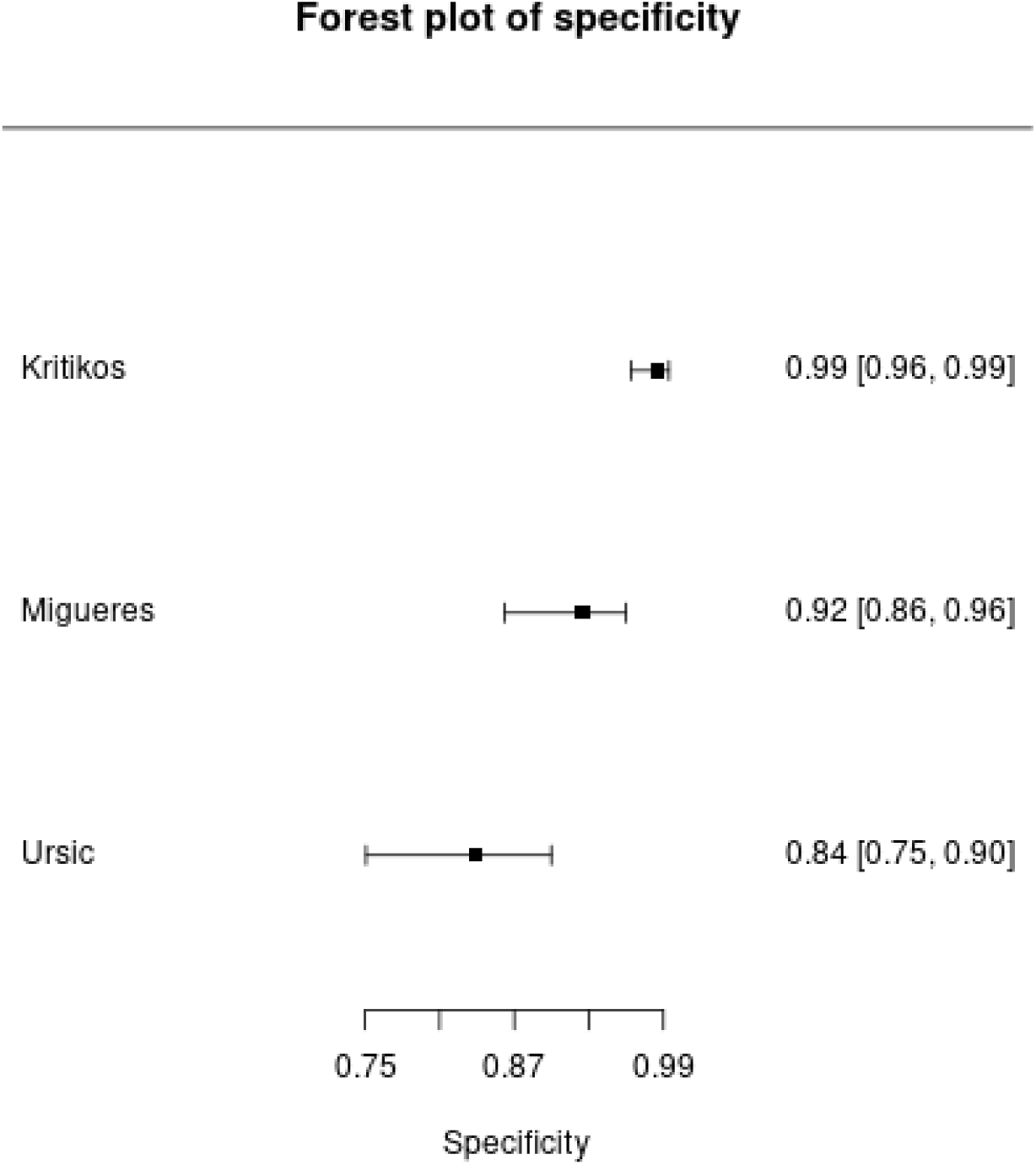
Forest plot of specificity for saliva versus nasopharyngeal sampling.

The only study investigating AN versus NP sampling (202 patients) showed a sensitivity of 90% (95% CI 82% to 95%) and a specificity of 99% (95% CI 95% to 100%).

### Certainty of evidence

Certainty of evidence for sensitivity and specificity pooled across studies were assessed and the absolute effect was interpreted per 100,000 patients with a pre-test probability of 0.0279%.

In the analysis of saliva versus NP sampling certainty of evidence was assessed as low-certainty of evidence for sensitivity. The assessment included three studies with 488 patients. Risk of bias and inconsistency were each downgraded one level. Risk of bias was downgraded one level because all included studies were assessed as unclear risk of bias. Inconsistency was downgraded one level because of non-overlapping 95% CIs.

For specificity, the certainty of evidence was assessed as very low-certainty of evidence. This assessment included three studies with 493 patients. Risk of bias and inconsistency were downgraded one level similar to the assessment of sensitivity. Imprecision was also downgraded one level.

If we have a pre-test probability of having SARS-CoV-2 infection of 0.0279%, 5,824 people out of 100,000 people would have positive test results. 26 of these positive test results would be true positives and 5,798 would be false positives. In contrast, 94,176 people out of 100,000 people would have a negative test result. Of these negative test results, 94,174 test results would be true negatives and 2 test results would be false negatives (Supplemental Material Figure S1).

In the analysis of AN sampling versus NP sampling certainty of evidence was assessed as very low for sensitivity and specificity. Both outcomes include only one single study with 202 patients. The effect was assessed per 100,000 patients with a pre-test probability of 0.0279%. Risk of bias was downgraded one level because a cut-off Ct-value and the sequence of sampling have not been reported. Imprecision was downgraded one level because of inclusion of very few participants (single study).

Thus, if we have a pre-test probability of having SARS-CoV-2 infection of 0.0279%, 1,025 per 100,000 people would have a positive test result. Of these positive test results, 25 would be true positives and 1,000 would be false positives. 98,975 people out of 100,000 people would have negative test results. 98,972 would be true negative test results and 3 would be false negative test results (Supplemental Material Figure S2).

## 4 DISCUSSION

To the best of our knowledge, this is the first systematic review with meta-analysis investigating the impact of sampling site on test accuracy using RT-PCR for the diagnosis of SARS-CoV-2 infection since the emergence of omicron.

After reviewing three DTA studies, we found saliva sampling to be 92% (95% CI 87% to 96%) sensitive and 94% (95% CI 83% to 98%) specific in detecting SARS-CoV-2 compared to NP sampling. All included studies did not report Ct-cut off values of positivity or the sequence of sampling performance. This impacted the assessment of certainty of evidence for sensitivity. Even the inconsistency between the single studies was high which is reflected in the non-overlapping 95% CIs leading to a further downgrading of the certainty of evidence. This inconsistency could be explained by different sampling techniques, different RT-PCR kits used, different sampling time points as well as a variable time point of symptom onset. However, due to the limited number of included studies subgroup analysis considering effect modifiers was not possible.

The certainty of evidence for specificity is further downgraded as the recommended threshold of 97% is not obtained. Therefore, the overall assessment of the certainty of evidence is only low for sensitivity und very low for specificity. This means that further research is very likely to have an important impact on our confidence in the estimate of effect and is likely to change the estimate. The high risk of becoming an incorrect diagnosis is shown in the absolute numbers of TP, FP, TN and FN at a pre-test probability of 0.0279%. Out of 100,000 people, only 26 people will have a true positive test result with saliva sampling. In contrast, 5798 people would have a false positive test result leading to incorrect diagnosis.

The same applies for the analysis of the single study for AN sampling versus NP sampling. In this study, we found AN sampling to be 90% (95% CI 82% to 95%) sensitive and 99% (95% CI 95% to 100%) specific. The certainty of evidence was assessed as very low for sensitivity and specificity. This means that any estimate of effect is very uncertain. The absolute numbers of TP, FP, TN and FN at a pre-test probability of 0.0279% pointed up the high risk for an incorrect diagnosis. Out of 100,000 people, only 25 people will have a true positive test result. On the contrary, 1,000 people will have a false positive test result.

Overall, the number of included patients per study is low and even the design of the studies and the patient setting is quite heterogeneous. Only one study focuses specifically on health care workers (HCWs), all other studies analyse the general population. Although we assume similar results between general population and HCWs in diagnostic test results this has to be mentioned. Even the different sampling methods of saliva could have an impact on our analysis regarding sensitivity and specificity.

Recent studies describe saliva sampling as a reliable tool in detecting SARS-CoV-2^24,25^ although these studies were performed before the emergence of the Omicron variant. Most studies comparing different sampling sites use antigen-rapid diagnostic tests (Ag-RDTs) and are reporting an unusually low sensitivity for saliva samples^26–29^. This is in concordance with our findings that saliva and AN sampling do not reach comparable sensitivity to NP sampling.

The Infectious Diseases Society of America (IDSA) also reviewed the published literature up to October 2020 to assess the performance of different specimen types relative to NP sampling for the detection of SARS-CoV-2^30^. For saliva sampling without coughing (9 studies, 387 patients) or with coughing (3 studies, 137 patients) the authors found a sensitivity of 90% and 99%, and a specificity of 98% and 96%, all assessed with low-certainty evidence. For AN sampling they reported a sensitivity of 89% and a specificity of 100%. Oropharyngeal sampling showed the lowest sensitivity of 76% with a specificity of 98%. The IDSA panel suggests collecting a NP swab, mid-turbinate swab, AN swab, saliva or a combined AN/oropharyngeal swab rather than an oropharyngeal swab alone for SARS-CoV-2 RNA testing in symptomatic individuals suspected of having COVID-19 (conditional recommendation, very low-certainty of evidence)^30^. Although the analysis of the IDSA was done before the Omicron period, the estimated effects are similar to our analysis. However, the guideline interpreted the effect with regard to a pre-test probability of 10% for symptomatic COVID-19 patients. In contrast, we interpret our findings in the context of the pre-test probability of 0.0279%, which corresponds to SARS-CoV-2 infections in May 2023.

Overall, saliva is a quite complex sample matrix, particularly if sputum or mucus is mixed with the sample. Even coughing or not coughing before the sample collection can make a difference in the analysis of the sample.

In summary, additional studies investigating non-invasive specimen-types such as saliva or AN are needed, even in pediatric patients or asymptomatic individuals, to determine accuracy of this less invasive and more comfortable sample collection methods.

## 5 LIMITATIONS

There are important limitations which restrict the interpretation and transferability of the findings. For example, the included studies differ in patients characteristics (e.g., symptom status, sampling time) and methodology of RT-PCR. However, due to the low number of studies we were not able to perform subgroup analyses with regard to important effect modifiers (e.g., symptom status). Therefore, there is uncertainty whether there might be different effects in populations of symptomatic or asymptomatic SARS-CoV-2 infected or uninfected people.

## 6 CONCLUDING REMARKS

In our analysis, both saliva sampling and AN sampling reach not comparable sensitivities and specificities as NP sampling. If there exist specific indications for using another sampling site than NP, it should be taken into account that this could lead to an increased number of false positive and negative results according to our findings. However, due to the low number of included studies we are not able to distinguish scenarios with asymptomatic and symptomatic infected people, and uninfected people. Due to the very low- to low-certainty of evidence, we are uncertain about the accuracy of saliva and AN sampling for diagnosis of SARS-CoV-2 compared to NP sampling.

## AUTHOR CONTRIBUTIONS

Conception of the review (SW); design of the review (SW, LSB); co-ordination of the review (SW, LSB); search and selection of studies for inclusion in the review (SW, LSB, NS, CW, TD, AF, HJ, AB); collection of data for the review (SW, LSB, CW, NS, TD, AF, AB); assessment of the risk of bias in the included studies (SW, LSB, CW, TD, AF, AB, NS); analysis of data (SW, LSB, CW, TD, AF, AB, NS); assessment of the certainty in the body of evidence (SW, LSB, CW, TD, AF, AB, NS); interpretation of data (SW, LSB, CW, TD, AF, AB, NS), and writing of the review (SW, LSB, CW, TD, AF, AB, HJ, NS), review and editing of the review (SW, LSB, CW, TD, AF, AB, HJ, NS, SSt).

## ETHICAL STATEMENT

No ethical approval is needed as systematic reviewing is based on published trial results.

## DATA AVAILABILITY STATEMENT

All data relevant to the study are included in the article, uploaded as supporting information, or are available from the corresponding author on reasonable request.

## ABBREVATIONS

PHEIC: public health emergency of international concern
DTA: diagnostic test accuracy
HCWs: health care workers
NP: nasopharyngeal
PPV: positive predictive value
NPV: negative predictive value
TP: true positives
TN: true negatives
FP: false positives
FN: false negatives
Ct: cycle threshold
Grading of Recommendations, Assessment, Development and Evaluation: GRADE

## ACKNOWLEDGMENTS

We thank the Federal Ministry of Education and Research, Germany NUM 2.0 (funding number: 01KX2121); part of the project “PREPARED” for supporting this work. We thank Maria-Inti Metzendorf for peer-reviewing the search strategy. We thank Angela Kunzler, Saskia Lindner, Julia Dormann, Annika Ziegler, Karolina Dahms and Kelly Ansems for literature screening.

