## Supplemental Material for "Impact of sampling site on diagnostic test accuracy of RT-PCR in diagnosing Severe Acute Respiratory Syndrome Coronavirus 2 (SARS-CoV-2) infection since the emergence of omicron: a systematic review and meta-analysis"

**Table S1.** Full Search Strategy for Electronic Databases

X. 1. Search for Evidence Syntheses (DTA)

**PubMed (Date of search: 07/03/2023)**

#1
"COVID-19"[mh] OR coronavir*[tiab] OR coronovir*[tiab] OR "corona virus"[tiab] OR COVID[tiab] OR COVID-19[tiab] OR COVID19[tiab] OR ncov[tiab] OR n-cov[tiab] OR 2019nCoV[tiab] OR nCoV2019[tiab] OR betacoronavirus*[tiab] OR WN-CoV[tiab] OR WNCoV[tiab] OR HCoV-19[tiab] OR HCoV19[tiab] OR "2019 novel*"[tiab] OR "2019 nCoV"[tiab] OR 2019-ncov[tiab] OR SARS-CoV-2[tiab] OR SARSCoV-2[tiab] OR SARSCoV2[tiab] OR SARS-CoV2[tiab] OR "SARS-CoV 2"[tiab] OR "SARS Cov"[tiab] OR sarscov[tiab] OR SARSCov19[tiab] OR SARS-Cov19[tiab] OR SARSCov-19[tiab] OR SARS-Cov-19[tiab] OR Ncovor[tiab] OR Ncorona*[tiab] OR Ncorono*[tiab] OR NcovWuhan*[tiab] OR NcovHubei*[tiab] OR NcovChina*[tiab] OR NcovChinese*[tiab] OR SARS2[tiab] OR SARS-2[tiab] OR "SARS 2"[tiab] OR "SARS 2"[tiab]

#2
"COVID-19 Testing"[mh] OR "antigen test*"[tiab] OR "ag test*"[tiab] OR "swab test*"[tiab] OR "nasal swab*"[tiab] OR "nasal test*"[tiab] OR "nasopharyngeal swab*"[tiab] OR "self test*"[tiab] OR "antigen detection*"[tiab] OR C19ST[tiab] OR "diagnostic test*"[tiab] OR "point of care"[tiab] OR "lateral flow"[tiab] OR RDT[tiab] OR RDTs[tiab] OR bedside[tiab] OR LFA[tiab] OR "polymerase chain reaction"[tiab] OR PCR[tiab] OR PCRs[tiab] OR qPCR[tiab] OR qPCRs[tiab] OR "RNA-directed DNA Polymerase"[tiab] OR "reverse-transcription loop-mediated isothermal amplification"[tiab] OR RT-LAMP[tiab] OR "nucleic acid test*"[tiab] OR POCT[tiab] OR POCTs[tiab] OR "NAAT"[tiab] OR "rapid nucleic acid amplification test*"[tiab] OR "rapid test*"[tiab] OR "rapid detection*"[tiab] OR RALFT[tiab]

#3
("Sensitivity and Specificity"[mh:noexp]) OR "Diagnostic Techniques and Procedures"[mh] OR DTA[tiab] OR diagnos*[tiab] OR detect*[tiab] OR casefinding[tiab] OR "case finding"[tiab] OR screen*[tiab] OR sensitivity[tiab] OR specificity[tiab] OR accuracy[tiab]

#4 systematic[sb]

#5
#1 AND #2 AND #3 AND #4

#6 Time Span: 01/01/2022 – 07/02/2023

#5 AND #6 (160)

**Web of Science (Clarivate) (Date of search: 07/02/2023)**

**Science Citation Index Expanded (1945-present)**

#1
(TI=((coronavir* OR coronovir* OR (coron* NEAR/2 (virus* OR viral* OR virinae*)) OR COVID OR COVID-19 OR COVID19 OR ncov OR n-cov OR 2019nCoV OR nCoV2019 OR betacoronavirus* OR WN-CoV OR WNCoV OR HCoV-19 OR HCoV19 OR "2019 novel*" OR "2019 nCoV" OR 2019-ncov OR 2019nCoV OR SARS-CoV-2 OR SARSCoV-2 OR SARSCoV2 OR SARS-CoV2 OR "SARS-CoV 2" OR "SARS Cov" OR sarscov OR SARSCov19 OR SARS-Cov19 OR SARSCov-19 OR SARS-Cov-19 OR Ncovor OR Ncorona* OR Ncorono* OR NcovWuhan* OR NcovHubei* OR NcovChina* OR NcovChinese* OR SARS2 OR SARS-2 OR "SARS 2"))) OR AB=(((coronavir* OR coronovir* OR (coron* NEAR/2 (virus* OR viral* OR virinae*)) OR COVID OR COVID-19 OR COVID19 OR ncov OR n-cov OR 2019nCoV OR nCoV2019 OR betacoronavirus* OR WN-CoV OR WNCoV OR HCoV-19 OR HCoV19 OR "2019 novel*" OR "2019 nCoV" OR 2019-ncov OR 2019nCoV OR SARS-CoV-2 OR SARSCoV-2 OR SARSCoV2 OR SARS-CoV2 OR "SARS-CoV 2" OR "SARS Cov" OR sarscov OR SARSCov19 OR SARS-Cov19 OR SARSCov-19 OR SARS-Cov-19 OR Ncovor OR Ncorona* OR Ncorono* OR NcovWuhan* OR NcovHubei* OR NcovChina* OR NcovChinese* OR SARS2 OR SARS-2 OR "SARS 2")))

#2
(TI=("antigen test*" or "ag test*" or ag-test* or "swab test*" or "nasal swab*" or "nasal test*" or "nasopharyngeal swab*" or self-test* or "self test*" or "antigen detection*" or C19ST or "diagnostic test*" or point-of-care or "point of care" or "lateral flow" or RDT or RDTs or bedside or LFA or Ag-RDT or Ag-RDTs or "polymerase chain reaction" or PCR or PCRs or RT-qPCR or RT-qPCRs or RT-PCR or RT-PCRs or "reverse-transcription-PCR" or "reverse-transcriptase-polymerase chain reaction" or rRT-PCR or rRT-PCRs or "RNA-directed DNA Polymerase" or "reverse-transcription loop-mediated isothermal amplification" or RT-LAMP or "nucleic acid test*" or POCT or POCTs or NAAT or "rapid nucleic-acid-amplification-test*" or "rapid nucleic acid amplification test*" or "rapid test*" or "rapid detection*" or RALFT)) OR AB=(("antigen test*" or "ag test*" or ag-test* or "swab test*" or "nasal swab*" or "nasal test*" or "nasopharyngeal swab*" or self-test* or "self test*" or "antigen detection*" or C19ST or "diagnostic test*" or point-of-care or "point of care" or "lateral flow" or RDT or RDTs or bedside OR LFA or Ag-RDT or Ag-RDTs or "polymerase chain reaction" or PCR or PCRs or RT-qPCR or RT-qPCRs or RT-PCR or RT-PCRs or rCT-PCR or rCT-PCRs or "RNA-directed DNA Polymerase" or "reverse-transcription loop-mediated isothermal amplification" or RT-LAMP or "nucleic acid test*" or POCT or POCTs or NAAT or "rapid nucleic-acid-amplification-test*" or "rapid nucleic acid amplification test*" or "rapid test*" or "rapid detection*" or RALFT))

#3
(TI=(DTA or diagnos* or detect* or screen* or casefinding or "case finding")) OR AB=(DTA or diagnos* or detect* or screen* or casefinding or "case finding")

#4
#1 AND #2 AND #3 (13'257)

#5
TI=("systematic review" or metaanalys* or meta-analys*) OR AB=("systematic review" or metaanalys* or meta-analys*)

#6
#3 AND #4 AND #5

Time span: 01/01/2022 – 07/02/2023
(107)

Total number of evidence syntheses retrieved: 267

After deduplication: 181

-----------------------------------------------------------------------------------------------------------------------------

2. Search for primary studies (DTA and non-DTA studies)

Study designs: Randomized controlled trials, cohort studies, cross-sectional studies, controlled-before-after studies

**PubMed (Date of search: 07/02/2023)**

#1
"COVID-19"[mh] OR coronavir*[tiab] OR coronovir*[tiab] OR "corona virus"[tiab] OR COVID[tiab] OR COVID-19[tiab] OR COVID19[tiab] OR ncov[tiab] OR n-cov[tiab] OR 2019nCoV[tiab] OR nCoV2019[tiab] OR betacoronavirus*[tiab] OR WN-CoV[tiab] OR WNCoV[tiab] OR HCoV-19[tiab] OR HCoV19[tiab] OR "2019 novel*"[tiab] OR "2019 nCoV"[tiab] OR 2019-ncov[tiab] OR SARS-CoV-2[tiab] OR SARSCoV-2[tiab] OR SARSCoV2[tiab] OR SARS-CoV2[tiab] OR "SARS-CoV 2"[tiab] OR "SARS Cov"[tiab] OR sarscov[tiab] OR SARSCov19[tiab] OR SARS-Cov19[tiab] OR SARSCov-19[tiab] OR SARS-Cov-19[tiab] OR Ncovor[tiab] OR Ncorona*[tiab] OR Ncorono*[tiab] OR NcovWuhan*[tiab] OR NcovHubei*[tiab] OR NcovChina*[tiab] OR NcovChinese*[tiab] OR SARS2[tiab] OR SARS-2[tiab] OR "SARS 2"[tiab] OR "SARS 2"[tiab]

#2
"COVID-19 Testing"[mh]"antigen test*"[tiab] OR "ag test*"[tiab] OR ag-test*[tiab] OR "swab test*"[tiab] OR "nasal swab*"[tiab] OR "nasal test*"[tiab] OR "nasopharyngeal swab*"[tiab] OR self-test*[tiab] OR "self test*"[tiab] OR "antigen detection*"[tiab] OR C19ST[tiab] OR "diagnostic test*"[tiab] OR point-of-care[tiab] OR "point of care"[tiab] OR "lateral flow"[tiab] OR RDT[tiab] OR Ag-RDT[tiab] OR Ag-RDTs[tiab] OR "polymerase chain reaction"[tiab] OR PCR[tiab] OR PCRs[tiab] OR RT-qPCR[tiab] OR RT-qPCRs[tiab] OR RT-PCR[tiab] OR RT-PCRs[tiab] OR "reverse-transcription-PCR"[tiab] OR "reverse-transcriptase-polymerase chain reaction"[tiab] OR rRT-PCR[tiab] OR rRT-PCRs[tiab] OR "RNA-directed DNA Polymerase"[tiab] OR "reverse-transcription loop-mediated isothermal amplification"[tiab] OR RT-LAMP[tiab] OR "nucleic acid test*"[tiab] OR POCT[tiab] OR POCTs[tiab] OR "rapid NAAT"[tiab] OR "rapid nucleic-acid-amplification-test*"[tiab] OR "rapid nucleic acid amplification test*"[tiab] OR "rapid test*"[tiab] OR "rapid detection*"[tiab] OR RALFT[tiab]

#3
("Sensitivity and Specificity"[mh:noexp]) OR "Diagnostic Techniques and Procedures"[mh] OR DTA[tiab] OR diagnos*[tiab] OR detect*[tiab] OR casefinding[tiab] OR "case finding"[tiab]

#4
(randomized controlled trial[pt] OR controlled clinical trial[pt] OR randomized[tiab] OR placebo[tiab] OR drug therapy[sh] OR randomly[tiab] OR trial[tiab] OR groups[tiab]) NOT (animals[mh] NOT humans[mh])

#5
"Cohort Studies"[mh:noexp] OR "Prospective Studies"[mh] OR "Retrospective Studies"[mh] OR "cohort stud*"[tiab] OR "prospective stud*"[tiab] OR "retrospective stud*"[tiab]

#6
#4 OR #5

#7
#1 AND #2 AND #3 AND #6
(3'858)

**Cochrane COVID-19 Study Register (https://covid-19.cochrane.org)
(Date of search for DTA studies: 07/02/2023 / Date of search for non-DTA studies: 09/03/2023)
(includes: PubMed, Embase, CENTRAL, ClinicalTrials.gov, WHO ICTRP, medRxiv, RetractionWatch)**

#1
"antigen test" OR "antigen tests" OR "ag testing" OR "ag test" OR "ag tests" OR "self test" OR "self tests" OR "antigen detection" OR "swab testing" OR "swab test" OR "swab tests" OR "nasal swab" OR "nasal swabs" OR "nasal test" OR "nasal tests" OR "nasopharyngeal swab" OR "nasopharyngeal swabs" OR C19ST OR "diagnostic test" OR "diagnostic tests" OR "point of care" OR "lateral flow" OR RDT OR RDTs OR "polymerase chain reaction" OR PCR OR PCRs OR qPCR OR qPCRs OR "reverse transcriptase polymerase" OR "polymerase chain reaction" OR "RNA directed DNA Polymerase" OR "reverse transcription loop mediated isothermal amplification" OR "RT LAMP" OR "nucleic acid test" OR "nucleic acid tests" OR POCT OR POCTs OR "rapid NAAT" OR "rapid nucleic acid amplification test" OR "rapid nucleic acid amplification tests" OR "rapid test" OR "rapid tests" OR "rapid detection" OR RALFT

#2
DTA or diagnos* or detect* or casefinding or "case finding" OR sensitivity OR specificity OR accuracy

#3 created: 1 Jan '22 - 7 Feb '23

#4
4a: Study design: Case Series/Case Control/Cohort

4b: Study design: Cross-sectional

4c: Randomized

#5 (DTA)
#1 AND #2 AND #3 AND #4
(3'744)

#6 (non-DTA)
#1 AND #3 AND #4
(8'354)

**WHO COVID-19 Research Database (search.bvsalud.org/global-literature-on-novel-coronavirus-2019-ncov/) (Date of search for DTA studies: 07/02/2023 / Date of search for non-DTA studies: 09/03/2023)**

**Databases selected**: PREPRINT-MEDRXIV, Scopus, PREPRINT-RESEARCHSQUARE, Web of Science, EuropePMC, ProQuest Central OR Academic Search Complete

Title, abstract, subject: ("antigen test" OR "antigen tests" OR "ag testing" OR "ag test" OR "ag tests" OR "self test" OR "self tests" OR "antigen detection" OR "swab testing" OR "swab test" OR "swab tests" OR "nasal swab" OR "nasal swabs" OR "nasal test" OR "nasal tests" OR "nasopharyngeal swab" OR "nasopharyngeal swabs" OR c19st OR "diagnostic test" OR "diagnostic tests" OR "point of care test" OR "point of care tests" OR "lateral flow" OR "polymerase chain reaction" OR pcr OR pcrs OR qpcr OR qpcrs OR "reverse transcriptase polymerase" OR "polymerase chain reaction" OR "RNA directed DNA Polymerase" OR "reverse transcription loop mediated isothermal amplification" OR "RT LAMP" OR "nucleic acid test" OR "nucleic acid tests" OR "rapid NAAT" OR "rapid nucleic acid amplification test" OR "rapid nucleic acid amplification tests" OR "rapid test" OR "rapid tests" OR "rapid detection" OR ralft) AND (dta OR diagnos* OR detect* OR casefinding OR "case finding" OR sensitivity OR specificity OR accuracy) AND ((cohort* OR prospective OR retrospective OR "follow up" OR "cross sectional" OR (control AND (study OR group)) OR "comparative study" OR "clinical trial") OR (random* OR placebo OR trial OR groups)) AND db:("PREPRINT-MEDRXIV" OR "Scopus" OR "PREPRINT-RESEARCHSQUARE" OR "Web of Science" OR "EuropePMC" OR "ProQuest Central" OR "Academic Search Complete")

**TABLE S2.** QUADAS-2 assessment interpretation guide

**Domain 1: Patient Selection:**

A. Risk of Bias

Signaling question 1: Was a consecutive or random sample of patients enrolled?

We scored ‘yes’ if the study enrolled a consecutive or random sample of eligible patients; ‘no’ if the study selected patients by convenience, and ‘unclear’ if the study did not report the manner of patient selection or unable to tell.

Signaling question 2: Was a case-control design avoided?

We scored ‘no’ if the study selected samples with a known RT-PCR results. We scored ‘yes’ if the status of the samples was unknown. We scored ‘unclear’ if we could not tell.

Signaling question 3: Did the study avoid inappropriate exclusions?

We scored ’yes’ to studies which included all participants regardless of symptoms or duration of symptoms. We scored ’no’ if studies excluded participants on the basis of symptoms or duration of symptoms. We scored ’unclear’ if we could not tell.

- Could the selection of patients have introduced bias?

Risk of Bias was scored ‘low risk’ if studies score ‘yes’ on all the question, ‘unclear risk’ if questions are answered with ‘yes’ and ‘unclear’, ‘high risk’ if two or more questions are answered with ‘no’.

B. Concerns regarding applicability

- Are there concerns that the included patients and setting do not match the review question?

We scored ’low concern’ if the study was conducted in a routine practice setting. We scored ‘high concern’ if patients did not met inclusion criteria or the study was not conducted in a routine setting. We scored ‘unclear concern’ if we could not tell.

**Domain 2: Index Test (s):**

A. Risk of Bias

Signaling question 1: Were the index test results interpreted without knowledge of the results of the reference standard?

Blinding is irrelevant if a Ct cut-off for test-positivity was defined and reported in the publication. A RT-PCR result is considered an objective measure if a Ct threshold was defined. We answered ’yes’ if the threshold (Ct cut-off for test positivity) was pre-specified and reported. We answered ’no’ if the threshold was not pre-specified, and ’unclear’ if we could not tell if the index test results were interpreted without knowledge of the reference standard results (threshold not reported).

Signaling question 2: If a threshold was used, was it pre-specified?

We answered ’yes’ if the threshold (Ct cut-off for test positivity) was pre-specified and reported. We scored ’no’ if the threshold was not pre-specified, and ’unclear’ if we could not determine if the threshold was prespecified or not.

Signaling question 3: Was the conduct of the RT-PCR described and followed the manufacturer’s instructions?

We answered ‘yes’ if the conduct of the RT-PCR was described and followed the manufacturer’s instructions, ‘no’ if the conduct of the RT-PCR did not follow the manufacturer’s instructions and ‘unclear’ if it was was not described and we could not tell.

Signaling question 4: Were storage conditions before testing appropriate?

We answered ‘yes’ if the storage conditions were reported and appropriate, ‘no’ if the storage conditions were not appropriate, ‘unclear’ if it was not reported and we could not tell.

- Could the conduct or interpretation of the index test have introduced bias?

Risk of Bias was scored ‘low risk’ if studies score ‘yes’ on all the question, ‘unclear risk’ if questions are answered with ‘yes’ and ‘unclear’, ‘high risk’ if two or more questions are answered with ‘no’.

B. Concerns regarding applicability

- Is there concern that the index test, its conduct or its interpretation differ from the review question?

If index test methods vary from those specified in the review question, concerns about applicability may exist. We scored ’high concern’ if the test procedure or its interpretation was inconsistent with the manufacturer recommendations, ’low concern’ if the test procedure and its interpretation was consistent with the manufacturer recommendations, and ’unclear concern’ if we could not tell.

**Domain 3: Reference standard:**

A. Risk of Bias

Signaling question 1: Is the reference standard likely to correctly classify the target condition?

We scored ‘yes’ for all studies as RT-PCR is the considered routine standard for detecting SARS-CoV-2.

Signaling question 2: Were the reference standard results interpreted without knowledge of the results of the index test?

If same procedure for RT-PCR used for index test and reference standard, the same answer was used in both cases. Blinding is irrelevant if a Ct cut-off for test-positivity was defined and reported in the publication. A RT-PCR result is considered an objective measure if a Ct threshold was defined. We answered ’yes’ if the threshold (Ct cut-off for test positivity) was prespecified and reported. We answered ’no’ if the threshold was not prespecified, and ’unclear’ if we could not tell if the index test results were interpreted without knowledge of the reference standard results (threshold not reported).

Signaling question 3: If a threshold was used, was it pre-specified and reported?

If same procedure for RT-PCR used for index test and reference standard, the same answer was used in both cases. We answered ’yes’ if the threshold (Ct cut-off for test positivity) was pre-specified and reported. We scored ’no’ if the threshold was not pre-specified, and ’unclear’ if we could not determine if the threshold was pre-specified or not.

Signaling question 4: Was the conduct of the RT-PCR described and followed the manufacturer’s instructions?

If same procedure for RT-PCR used for index test and reference standard, the same answer was used in both cases. We answered ‘yes’ if the conduct of the RT-PCR was described and followed the manufacturer’s instructions, ‘no’ if the conduct of the RT-PCR did not follow the manufacturer’s instructions and ‘unclear’ if it was was not described and we could not tell.

Signaling question 5: Were storage conditions before testing appropriate?

If same procedure for RT-PCR used for index test and reference standard, the same answer was used in both cases. We answered ‘yes’ if the storage conditions were reported and appropriate, ‘no’ if the storage conditions were not appropriate, ‘unclear’ if it was not reported and we could not tell.

- Could the reference standard, its conduct or its interpretation have introduced bias?

Risk of Bias was scored ‘low risk’ if studies score ‘yes’ on all the question, ‘unclear risk’ if questions are answered with ‘yes’ and ‘unclear’, ‘high risk’ if two or more questions are answered with ‘no’.

B. Concerns regarding applicability

- Is there concern that the target condition as defined by the reference standard does not match the review question?

If reference test methods vary from those specified in the review question, concerns about applicability may exist. We scored ’high concern’ if the test procedure or its interpretation was inconsistent with the manufacturer recommendations, ’low concern’ if the test procedure and its interpretation was consistent with the manufacturer recommendations, and ’unclear concern’ if we could not tell.

**Domain 4: Flow and Timing:**

A. Risk of Bias

Signaling question 1: Was there an appropriate interval between index test and reference standard?

We answered ‘yes’ if the sampling was described as simultaneous, ‘no’ if the sampling was not simultaneous and ‘unclear’ if we could not tell.

Signaling question 2: Was the sequence of sampling appropriate?

We answered ‘yes’ if saliva/nasal sampling was the first sampling and the nasopharyngeal sampling was the second sampling. We answered ‘no’ if saliva/nasal sampling was not the first sampling and ‘unclear’ if we could not tell.

Signaling question 3: Did all patients receive a reference standard?

We answered ‘yes’ for all studies as all studies used RT-PCR as reference standard

Signaling question 4: Did patients receive the same reference standard?

We answered ‘yes’ for all studies that used the same RT-PCR for all samples and ‘no’ if the samples were analyzed by different types of RT-PCR. We scored ‘unclear’ if we could not tell the used RT-PCR.

Signaling question 5: Were all patients included in the analysis?

We answered ’yes’ if the whole population was included in the analysis or reasons for exclusion were reported and appropriate. We answered ‘no’ if samples were excluded without a given reason or exclusion was inappropriate. We answered ‘unclear’ if we could not tell.

- Could the patient flow have introduced bias?

Risk of Bias is scored ‘low risk’ if studies score ‘yes’ on all the question, ‘unclear risk’ if questions are answered with ‘yes’ and ‘unclear’, ‘high risk’ if two or more questions are answered with ‘no’.

**FIGURES**

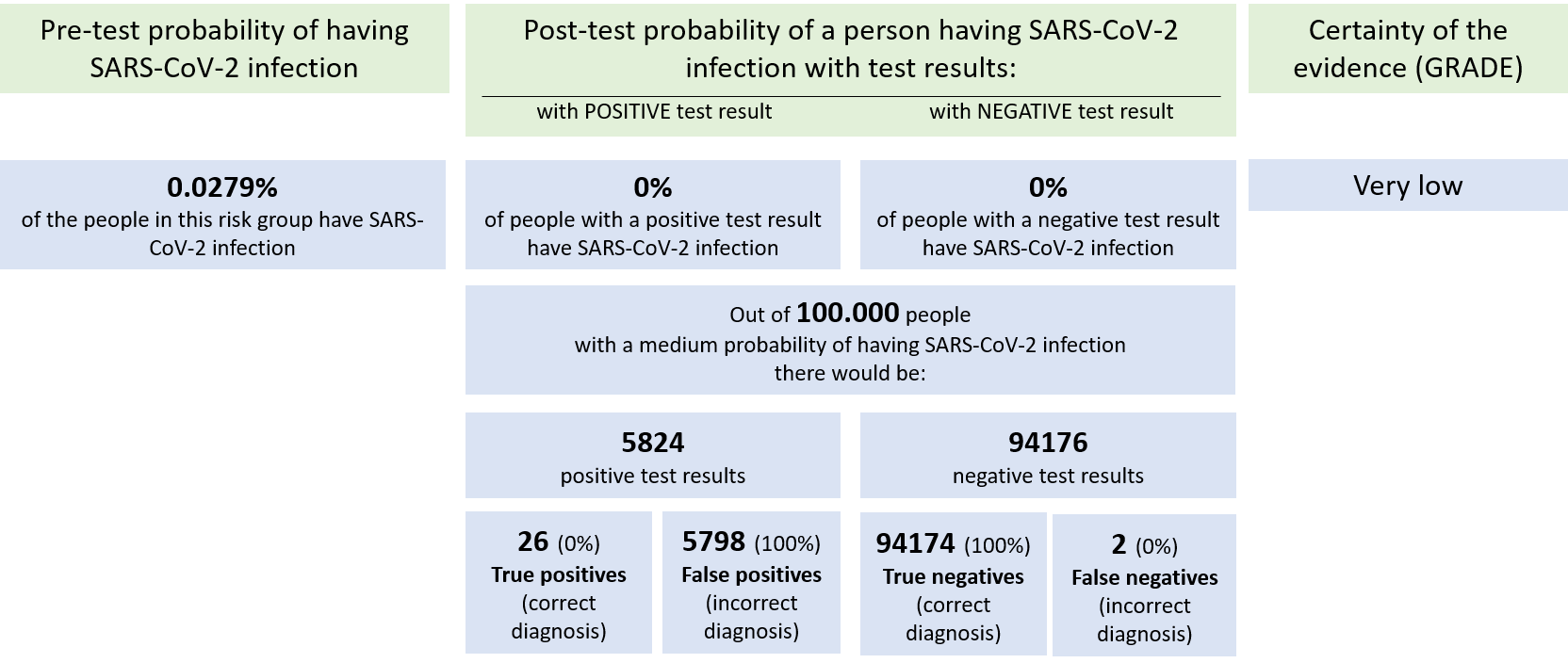

**Figure S1:** Summary of findings. Saliva sampling versus nasopharyngeal (NP) sampling. Modified from GRADEpro.

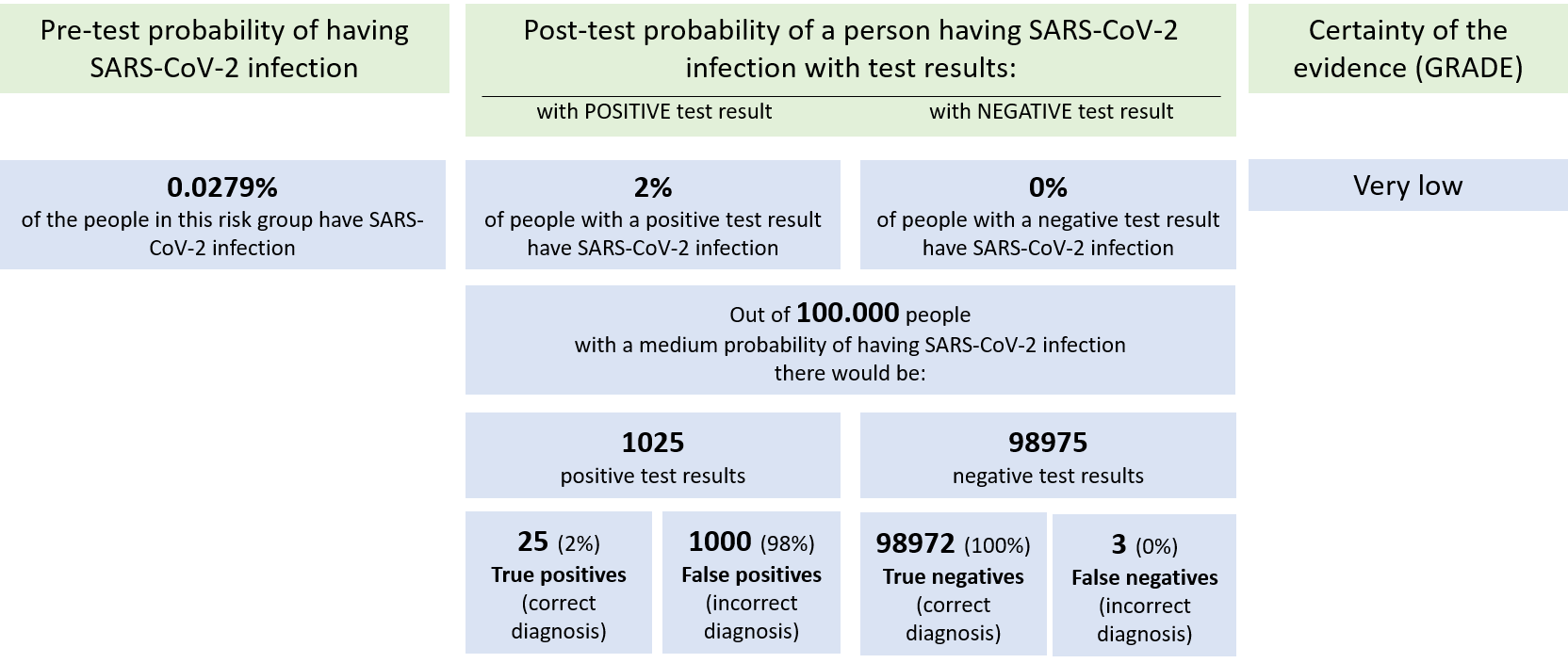

**Figure S2:** Summary of Findings. Anterior nasal (AN) sampling versus nasopharyngeal (NP) sampling. Modified from GRADEpro.
